# MEDIAL FRONTAL CORTEX GAMMA-AMINOBUTYRIC ACID CONCENTRATIONS IN PSYCHOSIS AND MOOD DISORDERS: A META-ANALYSIS OF PROTON MAGNETIC RESONANCE SPECTROSCOPY STUDIES

**DOI:** 10.1101/2022.02.21.22271287

**Authors:** Molly Simmonite, Clara J. Steeby, Stephan F. Taylor

**Author notes:** **Corresponding author:** Molly Simmonite.

## Abstract

**Background:** Abnormalities of gamma-aminobutyric acid-ergic (GABAegic) systems may play an important role in psychosis spectrum and mood disorders. Proton magnetic resonance spectroscopy allows for non-invasive in vivo quantification of GABA; however, studies of GABA in psychosis have yielded inconsistent findings. This may stem from grouping together disparate voxels from functionally heterogeneous regions.

**Methods:** We searched the PubMed database for magnetic resonance spectroscopy studies of medial frontal cortex (MFC) GABA in patients with psychosis, bipolar disorder, depression, and individuals meeting ultra-high risk for psychosis criteria. Voxel placements were classified as rostral-, rostral-mid-, mid-, or posterior MFC, and random effects meta-analyses conducted for each group, for each MFC sub-region.

**Results:** Of 341 screened articles, 23 studies of psychosis (752 patients,856 controls), 6 studies of bipolar disorder (129 patients, 94 controls), 20 studies of depression (463 cases, 449 controls) and 7 studies of ultra-high risk (229 patients, 232 controls) met inclusion criteria. Meta-analysis revealed lower mid-(SMD = -0.28, 95% confidence interval [CI] = -0.48 to -0.07, *p* < .01) and posterior (SMD = -0.29, 95% CI = -0.49 to -0.09, *p* <.01) MFC GABA in psychosis, and increased rostral GABA in bipolar disorder (SMD = 0.76, 95% CI = 0.25 to 1.25, *p* < .01). In depression, reduced rostral MFC GABA (SMD = -0.36, 95% CI = -0.64 to -0.08, *p* = .01) did not survive correction for multiple comparisons.

**Conclusions:** These results substantiate the relevance in the ethology of psychosis spectrum and mood disorders and underline the importance of voxel placement.

## INTRODUCTION

Substantial evidence from several lines of research has suggested that disturbances in the inhibitory neurotransmitter gamma-aminobutyric acid (GABA) play a role in the pathophysiology of both psychosis spectrum disorders and mood disorders. Consistent evidence from post-mortem and preclinical studies in schizophrenia and bipolar disorder reveal abnormalities in fast-spiking, parvalbumin-positive GABAergic interneurons (1), as well as reductions in mRNA and protein levels of the 67-kDa isoform of the GABA-synthesizing enzyme, glutamic acid decarboxylase (2–4). In depression, studies show reduced GABA concentrations in plasma (5,6) and cerebrospinal fluid (7–9), possibly linked to vulnerability of somatostatin-positive GABAergic interneurons (10).

Proton magnetic resonance spectroscopy (^1^H-MRS) is a powerful way to non-invasively investigate GABA concentrations *in-vivo*. Measuring GABA poses specific challenges due to its relatively low concentration and high spectral overlap with more abundant metabolites; however, sequences such as MEGA-PRESS (11,12) take advantage of couplings within the GABA molecule, which allow GABA signals to be reliably separated from stronger signals.

While the post-mortem evidence linking psychosis spectrum disorders to GABAergic disfunction is well-replicated, ^1^H-MRS studies have, thus far, yielded inconsistent findings. In schizophrenia for example, MRS studies have revealed increased (13,14) decreased (15,16) and as normal GABA concentrations (17), when patients are compared with controls. Attempts to reach consensus by pooling study data via meta-analysis also appear contradictory, with evidence for reduced GABA concentrations found in some analyses (18–20), but not others (21,22).

There are several possible reasons to explain these mixed findings. Individual studies may differ in the clinical characteristics of their samples, in terms of illness duration, symptom profile or medication use. Similarly, there may be large differences between studies in methodology, e.g. magnet strength, reference metabolites or tissue correction strategy used. Especially critical may be the location of the MRS voxel. Due to the relatively low concentration of GABA in comparison with other metabolites, large voxels, typically on the order of 3 × 3 × 3cm^3^, are collected to offset the low signal to noise ratio. The time to acquire these voxels is usually around 10 minutes per voxel, which means that researchers generally only have enough time to collect one or two voxels per study. When meta-analyses combine data from these studies, often voxels from disparate brain regions are combined – sometimes across the whole brain, or entire frontal cortex – sometimes with minimal overlap. Evidence in healthy controls however, indicates that GABA concentrations vary across regions (23–25).

The medial frontal cortex (MFC) is both functionally heterogeneous and strongly implicated in the etiologies of psychosis spectrum disorders and mood disorders. To map the functions of the subregions of the MFC, de la Vega and colleagues (26) used NeuroSynth (https://neurosynth.org/) to perform a meta-analysis which pooled the results of almost 10,000 fMRI studies. Their findings revealed distinct functional profiles in the MFC, with anterior MFC implicated in reward, decision making, social processing and episodic memory, the middle MFC with cognitive control, negative affect and pain, and the posterior in motor function. These regions likely contribute to symptoms of psychosis and depression in different ways; thus, combining GABA concentration findings across these functionally heterogeneous regions may not be the best strategy to gain the benefit of pooled studies in a meta-analysis.

Given the importance of the various functions of the MPFC for psychiatric disorders, as well as the number of studies that focus on the MPFC, we performed a meta-analyses of ^1^H-MRS studies of medial frontal GABA concentration in psychosis spectrum disorders, including individuals at an ultra-high risk (UHR), and depression. Our primary focus was on psychosis, given the relative strength of the post-mortem findings and focus of our prior work (27), and mood disorders were included for comparison. Voxels are classified as being in one of four medial frontal sub-regions – rostral, rostral-mid, mid, and posterior, and each of these sub-regions are examined separately. Our aim is to gain a more nuanced picture of the profile of medial frontal GABA dysfunction in psychiatric disorders.

## METHODS AND MATERIALS

### Search Strategy

Meta-analyses were conducted and reported according to the Preferred Reporting Items for Systematic Reviews ad Meta-Analyses (PRISMA) methodology. ^1^H-MRS studies that examined differences in GABA levels between healthy controls and patients with psychosis, bipolar disorder, depression, or individuals at ultra-high risk (UHR) for psychosis were obtained through a PubMed search, using the search term “(MRS OR Magnetic Resonance Spectroscopy) AND (GABA OR Gamma Aminobutyric Acid) AND (psychosis OR depression OR schizophrenia OR bipolar disorder). Databases were searched for articles published prior to October 25, 2021. Titles and abstracts were examined to determine suitability for inclusion in the analysis. Reference sections of the returned articles, as well as review articles (27) and meta-analyses (18,20,22,28–31) were also searched for additional articles that fulfilled our inclusion criteria.

Additionally, we also searched the online pre-print servers bioAriv, medRxiv and psyArXiv for studies which met our inclusion criteria. While the inclusion of “grey literature” such as preprints or dissertations may be controversial, analysis suggests that not including such literature in meta-analysis may result in the overrepresentation of studies with statistically significant results, and less precise effect sizes estimates (32).

### Study Selection

Studies met criteria for inclusion if they: (a) were an original research article; (b) used ^1^H-MRS to study *in vivo* GABA concentrations; (c) compared groups with psychosis, bipolar disorder, depression, or UHR, with healthy control participants; (d) included ^1^H-MRS voxels placed in the medial frontal cortex; (e) were published in, or translated into, the English language. Articles were screened for overlapping samples, and where present, data from the article reporting the largest sample was included. If an article presented GABA concentrations before and after an intervention, only baseline measures were included.

### Outcome measures

From each of the studies that met inclusion criteria, we extracted publication information (authors, year of publication), participant characteristics (diagnosis, sample size, age, gender, illness duration, medication status), methodological characteristics (field strength, acquisition sequence, reference metabolite), voxel location, and GABA concentration (mean and SD in each group). This information was extracted by one author (C.J.S.) and independently verified by another (M.S.). Where published articles did not include this information in the text, tables or supplementary materials, the authors were contacted, or where possible, values were estimated using an online tool (https://automeris.io/WebPlotDigitizer/).

### Meta-analysis

Voxels were categorized as rostral, mid, or posterior MFC. Where voxels straddled the rostral and mid regions of the MFC, they were classified as rostral-mid MFC. This classification was performed by one author (C.J. S.) and reviewed by the other authors (M.S. and S.F.T.). Separate meta-analyses were conducted for each MFC subregion (rostral, rostral-mid, mid, and posterior), for each clinical population (PSY, BD, DEP, and UHR).

Data were analyzed with R version 4.1.1, using the “metafor” package (33). The R code used to perform the meta-analyses is available in the supplementary materials. Meta-analysis was conducted where at least three datasets meeting inclusion criteria were available for a subregion for a clinical population of interest. Results were instead summarized if this number was not met. Psychosis samples were classified as acute (average duration of illness < 5 years) and chronic (average duration of illness ≥ 5 years), and depression samples were classified as depressed or remitted at the time of MRS scanning. Analyses of these subgroups were conducted if at least 5 datasets met inclusion criteria for that MFC subregion. Meta-regressions were performed to determine differences between the subgroups. Effect sizes were described using standardized mean differences (SMD; also known as Hedges’ g) and 95% confidence intervals. Use of SMD allowed for comparison of different units of GABA measurement (institutional units, ratios to reference metabolites). Random effects models were used to pool effect sizes, as we assumed heterogeneity in both the clinical profile of patient samples and the methodology employed in each study (e.g., magnet strengths, analysis software used, voxel tissue composition correction strategy). Since examining GABA concentrations in psychosis was our primary interest, we corrected for multiple comparisons by applying a Bonferroni-corrected threshold of *p* < 0.0125 (=0.05/4) to determine statistical significance, since we performed 4 independent meta-analyses. For the remaining meta-analyses of depression, bipolar disorder and UHR, we applied a Bonferroni-corrected threshold of *p* < 0.0083 (=0.05/6) for statistical significance, since we performed a total of 6 meta-analyses investigating these samples.

Between-study heterogeneity was assessed with the I^2^ index and Q-statistic. Higher I^2^ scores indicate higher variation between studies, with values of 25%, 50% and 75% representing small, moderate, and high levels of heterogeneity, respectively. Significant Q-statistics suggest the presence of heterogeneity but does not indicate the extent of this heterogeneity (34). Publication bias was assessed by visually examining funnel plots for asymmetry and performing Egger’s regression test for funnel plot asymmetry (35).

## RESULTS

### ^1^H-MRS study characteristics

The literature review identified a total of 54 studies matching our inclusion criteria. The PRISMA flow diagram is presented in Figure 1. Of the 54 studies, 23 studies included participants with psychosis (13– 15,17,36–53) (752 cases, 856 controls), 7 studies included individuals at UHR (47,53–58) (229 cases, 232 controls), 20 studies included individuals with depression (59–76) (463 cases, 499 controls), and 6 studies included participants with bipolar disorder (77–82) (129 cases, 94 controls). Two studies included multiple clinical samples (47,53), therefore these numbers sum 56. Detailed Study characteristics for are presented in Supplementary Tables 1-4. Voxels from each study were reviewed, and classified as rostral-, rostral-mid-, mid- and posterior MFC, with classifications shown in Figure 2.

**Figure 1:**
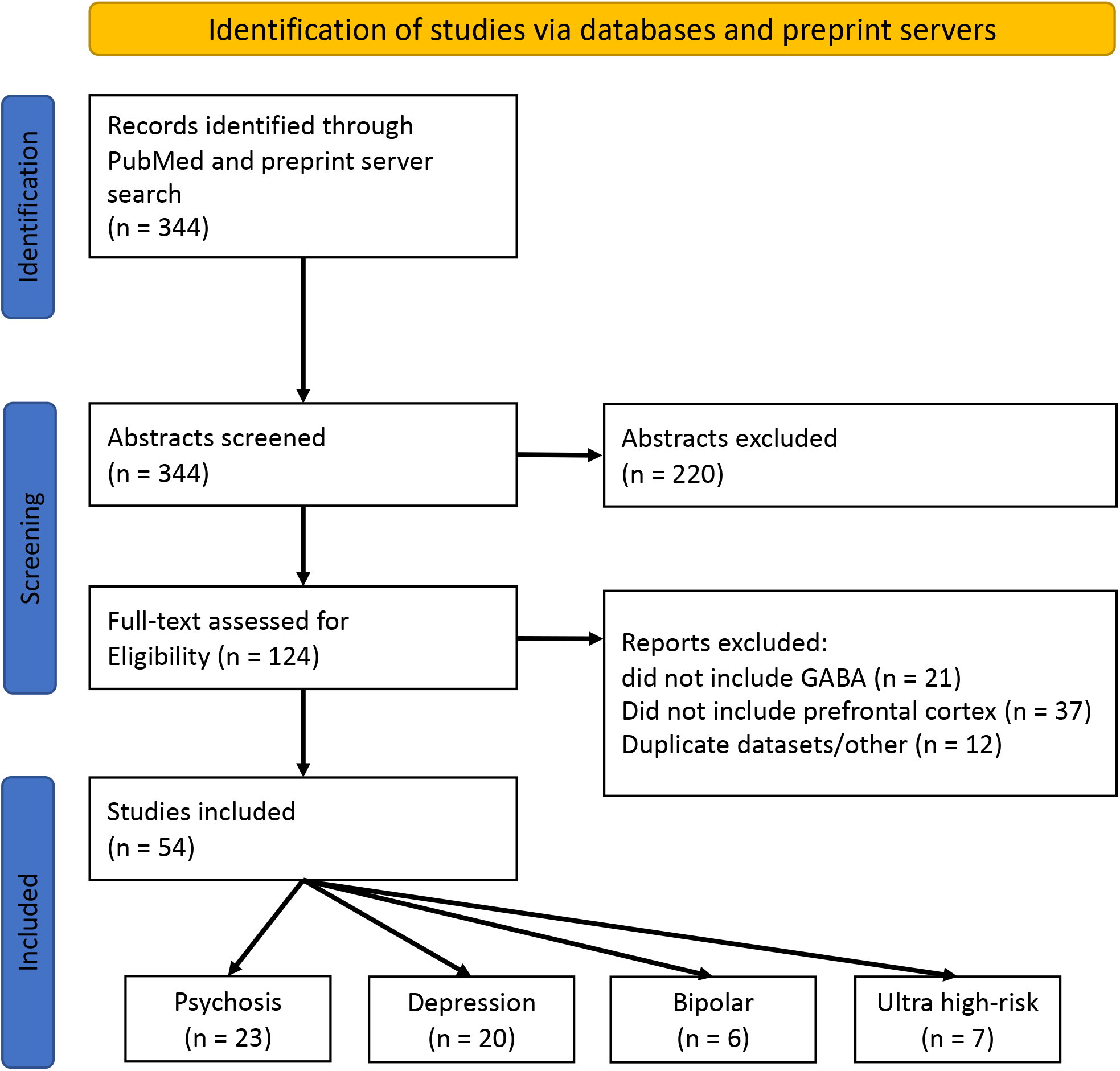
PRISMA flow diagram of search for meta-analyses. Note, two studies included both psychosis and ultra-high risk samples, and so the breakdown of included studies totals 56 due to these duplicates.

**Figure 2:**
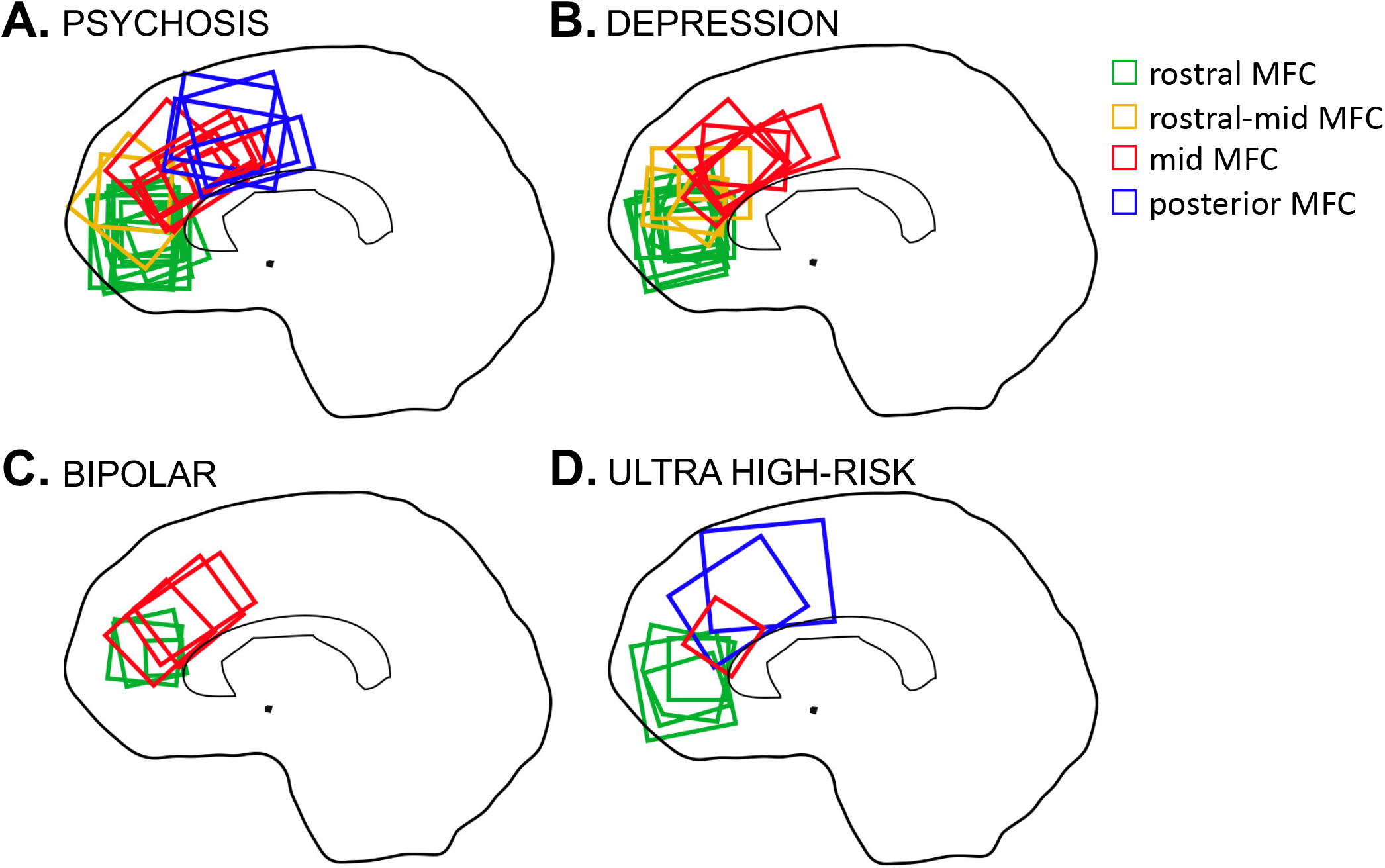
Voxel location in medial frontal cortex GABA ^1^H-MRS studies. A: ^1^H-MRS studies of psychosis; B: ^1^H-MRS studies of depression; C: ^1^H-MRS studies of bipolar disorder; and D: ^1^H-MRS studies of individuals meeting ultra-high risk of developing psychosis criteria.

### Medial Frontal GABA concentrations in Psychosis Spectrum and Mood Disorders

#### Psychosis

Figure 3 shows the results of the meta-analyses of individuals with psychosis. GABA concentrations were significantly reduced in both the mid-(SMD = -0.28, 95% CI = -0.48 to -0.07, *p* = .0087) and posterior MFC (SMD = -0.29, 95% CI = -0.49 to -0.09, *p* = .005) in patients with psychosis compared with healthy controls. I^2^ values were 36.50% and 0.00% respectively, suggesting small heterogeneity across studies. Sub-group analyses revealed that mid MFC GABA concentrations were significantly reduced in acute patients (SMD = -0.38, 95% CI = -0.58 to -0.17, *p* = 0.0003), but not chronic patients (SMD = -0.18, 95% CI = -0.58 to 0.23, *p* = .39); however meta-regression did not indicate significant differences between the subgroups. GABA concentrations in the rostral MFC showed no difference compared to controls (SMD = 0.11, 95% CI = -0.27 to 0.48, *p* = .58), and likewise no significant effects in the subgroup analyses of acute and chronic patients. This region was notable for high heterogeneity in the combined group (I^2^=74.83%), as well as acute (I^2^=79.10%) and chronic (I^2^=63.65%) subgroups. There were insufficient studies of posterior MFC GABA to perform subgroup analyses for this region. Reductions in GABA concentrations in the rostral-mid MFC in patients with psychosis did not survive corrections for multiple comparisons (SMD = -0.57, 95% CI = -1.10 to -0.03, *p* = .04).

**Figure 3:**
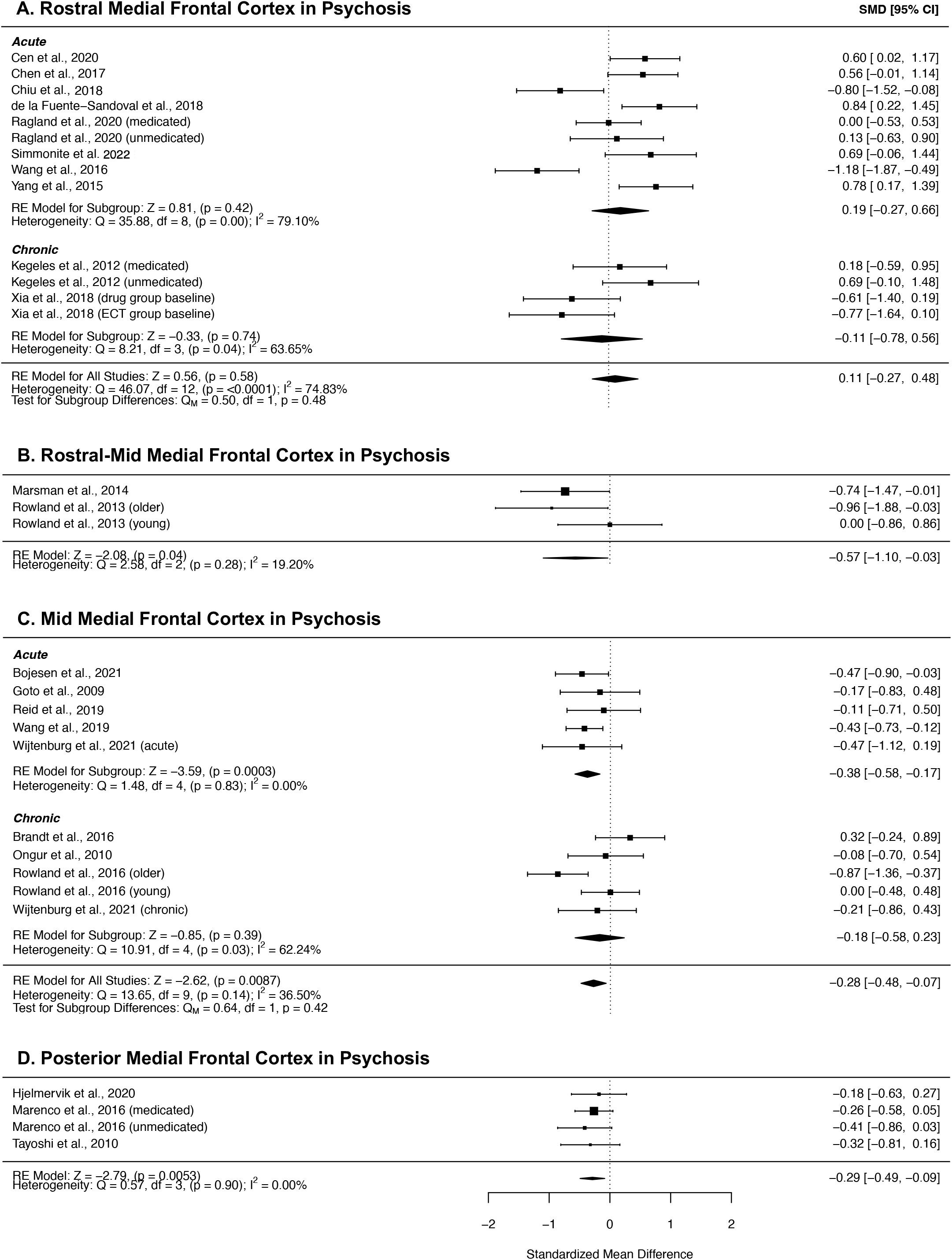
Forest plots showing summary effect sizes for group differences between individuals with psychosis and healthy controls in the A) rostral MFC; B) rostral-mid MFC, C) mid MFC and D) posterior MFC Negative SMDs denote lower GABA concentrations in patients than healthy controls; positive SMDs denote higher GABA concentrations in patients than healthy controls. Abbreviations: SMD – standardized mean difference, CI – confidence interval, RE – Random effects, df – degrees of freedom.

#### Depression

Forest plots detailing the results of the meta-analyses of GABA concentrations in studies investigating depression are presented in Figure 4. While the meta-analysis of rostral MFC GABA concentrations in depression indicated a reduction compared with controls, the significance of this effect did not survive correction for multiple comparisons (SMD = -0.36, 95% CI = -0.64 to -0.08, *p* = 0.01). Subgroup analyses of depressed patients revealed a similar effect size (SMD = -0.40, 95% CI = -0.70 to -0.11, *p* = 0.0073), but there were insufficient studies of remitted patients to perform an analysis of this group. Heterogeneity was moderate across the full sample (I^2^ = 48.15%) and within the depressed subgroup (I^2^ = 47.12%). We did not find any studies which reported posterior GABA concentrations in depression.

**Figure 4:**
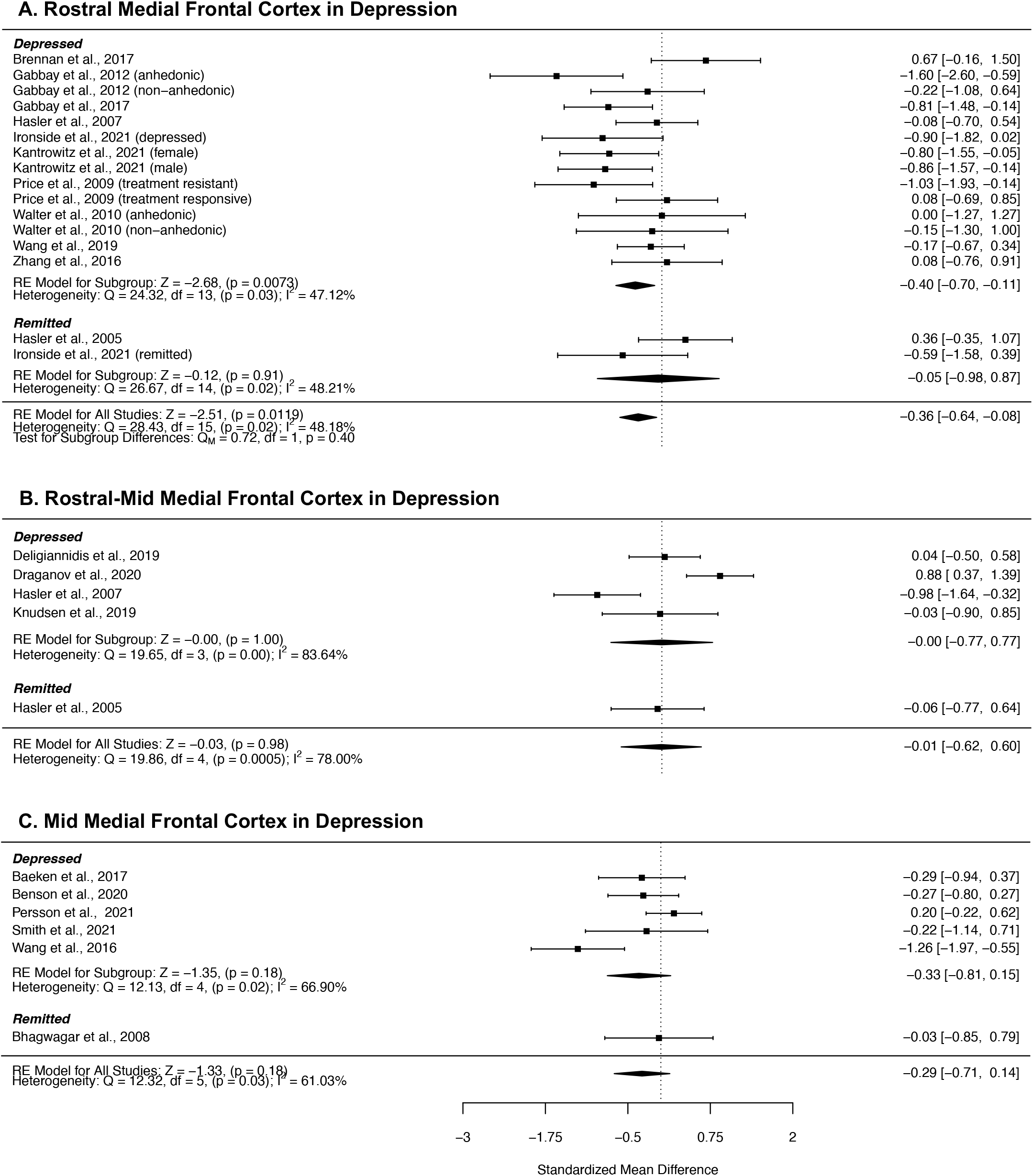
Forest plots showing summary effect sizes for group differences between individuals with depression and healthy controls in the A) rostral MFC, B) rostral-mid MFC and C, mid MFC. Negative SMDs denote lower GABA concentrations in patients than healthy controls; positive SMDs denote higher GABA concentrations in patients than healthy controls. Abbreviations: SMD – standardized mean difference, CI – confidence interval, RE – Random effects, df – degrees of freedom.

#### Bipolar Disorder

Figure 5a and 5b present the results of the meta-analyses of studies of bipolar disorder. GABA concentrations in rostral MFC were higher in patients with bipolar disorder compared with controls (SMD = 0.76, 95% CI = 0.26 to 1.25, *p* = 0.0026), and heterogeneity across studies was small (I^2^ = 0.00%). We did not find any studies which published analyses of posterior GABA.

**Figure 5:**
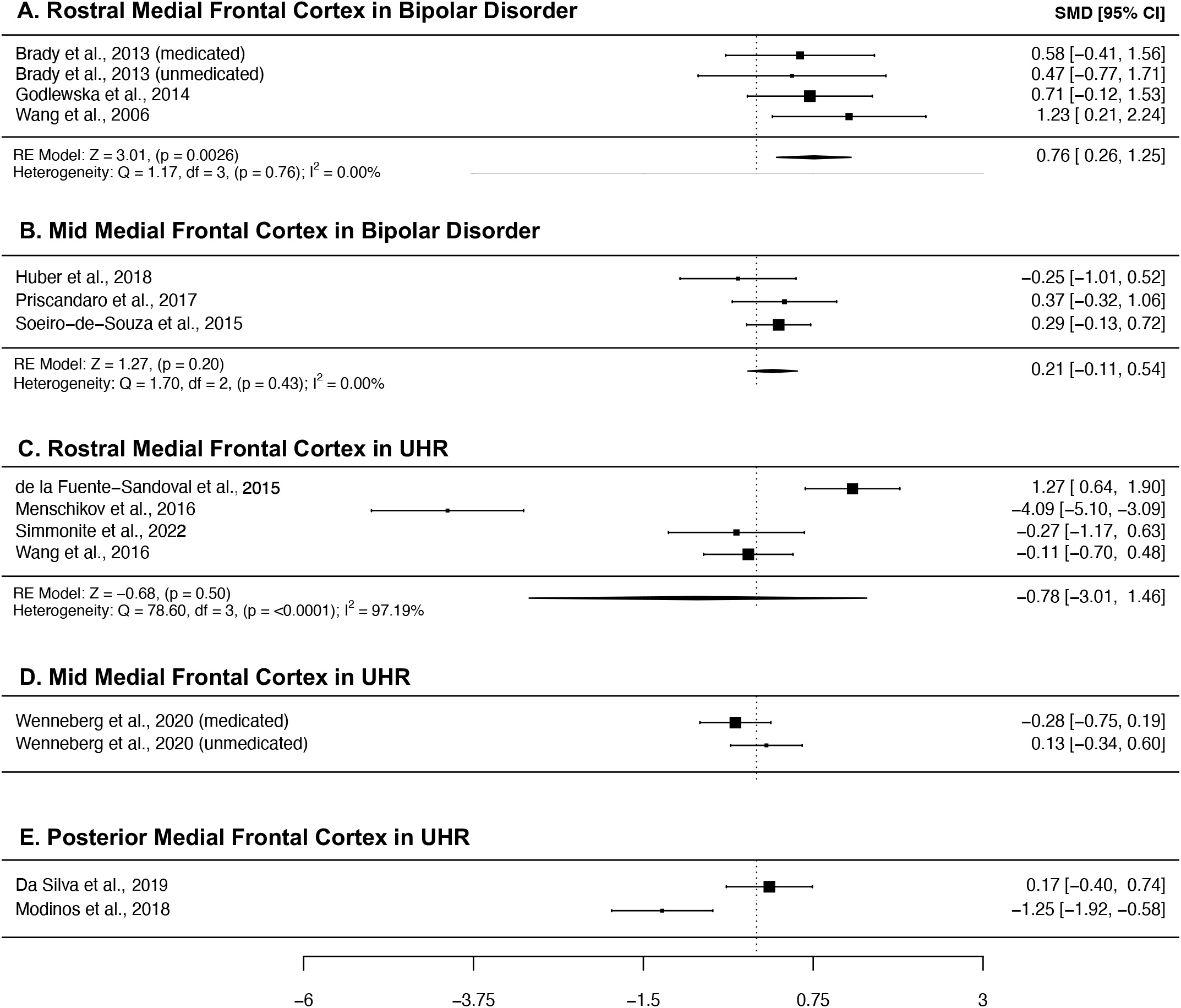
Forest plots showing summary effect sizes for group differences between individuals with bipolar disorder and healthy controls in the A) rostral MFC, and B) mid MFC, and individuals meeting ultra-high risk for psychosis criteria and healthy controls in the C) rostral MFC, D) mid MFC and E) posterior MFC. Negative SMDs denote lower GABA concentrations in patients than healthy controls, positive SMDs denote higher GABA concentrations in patients than healthy controls. Abbreviations: SMD – standardized mean difference, CI – confidence interval, RE – Random effects, df – degrees of freedom.

#### UHR

Plots summarizing the findings of ^1^H-MRS studies investigating GABA concentrations in UHR samples are presented in Figure 5c, 5d and 5e. Due to the limited number of studies published, we were only able to perform a meta-analysis of rostral MFC GABA, in which we found no significant differences between individuals at UHR and controls, and high between-study heterogeneity (I^2^ = 97.19%).

### Publication Bias

Visual inspection of funnel plots and results of Eggers test (provided in Supplementary Figures 1,2 and 3) did not suggest publication bias.

## DISCUSSION

The current study presents a series of meta-analyses in which regional GABA concentrations in the MFC are investigated, comparing individuals with psychosis spectrum disorders and depression with healthy controls. Our main findings were: (a) patients with psychosis have significantly decreased GABA+ concentrations in the mid- and posterior MFC (b) in patients with bipolar disorder we found Increased GABA+ concentrations in the rostral MFC (c) reduced GABA+ concentrations in the rostral MFC in depression did not survive correction for multiple comparisons.

### Decreased mid and posterior MFC GABA+ in psychosis

In individuals with psychosis, our analysis revealed significantly decreased GABA concentrations in the mid and posterior regions of the MFC, while there were no significant differences in the rostral, or rostral-mid MFC. Subgroup analysis revealed these declines were significant in acute, but not chronic patients, (i.e., those with an illness duration of greater than five years), suggesting that GABA abnormalities may be modulated by illness stage or mitigated by prolonged medication use. Our finding is similar to that of Nakahara et al. (19), who recently used meta-analyses to demonstrate midcingulate cortex (MCC), but not anterior cingulate cortex (ACC) GABA concentrations were reduced in in first episode psychosis (FEP) and a patient group comprised of FEP and schizophrenia patients, including an unmedicated subgroup. In contrast, Kumar et al.’s (18) investigation of frontal GABA in schizophrenia revealed reductions which were restricted to the ACC. Examination of the studies that included in their ACC region however, included substantial overlap with our posterior MFC region and Nakahara et al.’s (19) MCC region, which may explain this discrepancy.

Evidence from functional and structural studies have implicated the MFC as a key region in psychosis (83). It has been linked with a number of functions, including cognitive control (84), emotion regulation (85,86) and conflict monitoring (87,88), all of which are impaired in psychosis. While there has been debate about the precise functional mapping of the MFC, including the cingulate cortex, meta-analysis has implicated rostral regions of the MFC in reward, episodic memory and social processing, and more posterior regions in cognitive control (26). Our finding of decreased GABA concentrations in these regions are in line with post-mortem studies of schizophrenia, which have consistently shown reductions in the mRNA and protein levels of the 67-kDa isoform of GAD, which is responsible for the majority of GABA synthesis (2–4), as well as findings of impaired cognitive control in functional measures (89,90).

While we found evidence of GABA reduction in the mid and posterior MFC in individuals with psychosis, and our subgroup of acute psychosis, our search of the literature did not uncover enough investigations of GABA in these regions in UHR individuals for us to perform meta-analyses for that sample. Review of the available UHR publications does not suggest robust GABA reductions in either the mid-or posterior MFC, suggesting that the regional GABA reductions we found in patients with psychosis are associated with the onset of symptoms, rather than being a trait marker for vulnerability for psychosis.

### Increased rostral MFC GABA+ concentrations in bipolar disorder

Prior meta-analyses have found no significant differences in GABA+ concentrations, when including voxels from across the whole brain(20,28), or in the ACC(29). Our analysis focusing in on the MFC found significantly increased GABA+ concentrations in the rostral MFC. Increased GABA was not significant in the mid MFC and we were unable to perform an analysis of the posterior MFC, due to a lack of studies investigating this region. To our knowledge, we are the first meta-analysis of bipolar disorder to investigate GABA concentrations in subregions of the frontal cortex.

Our finding of significantly Increased GABA aligns with prior reports of elevated GABA levels in the plasma of bipolar patients(91,92). Increased GABA could be the result of a primary pathological process, or it could be a compensatory response, e.g., to environmental stressors. In rodents, a chronic unpredictable stress model was observed to increase GABA levels in the ACC, as measured by MRS(93). Increased GABA could also be a secondary response to glutamatergic dysfunction. Ketamine infusion, which blocks excitatory NMDA receptors, has been shown to increase mPFC GABA in humans(94).

The number of investigations of GABA+ concentrations in bipolar disorder available is limited, and therefore it is difficult to perform subgroup analyses based on medication status. Furthermore, often the samples reported are comprised of both medicated and unmedicated participants, which means meta-analyses are unable to untangle the effects of medication. One of the studies we included in our meta-analysis did consider the use of GABA modulating medications, such as benzodiazepines (which are often used to treat anxiety or insomnia in psychosis and mood disorders), and found that such medications partially correct GABA concentrations to a healthy control level in their sample(77). This finding indicates that medication may obscure GABA+ concentration increases in bipolar disorder.

### Decreased GABA+ in the rostral MFC in depression did not survive correction for multiple comparisons

Previous meta-analyses have revealed reduced GABA concentrations in depression when considering voxels from across the whole brain(20,30). When focusing on the frontal cortex, Schur et al.(20), found no evidence for abnormal GABA concentrations in depression, while Romeo et al., and Godfrey et al.(30), found significantly reduced GABA in the frontal cortex and ACC, respectively. While the studies included in Godfrey et al.’s(30) analysis of ACC GABA were a subsection of those included in our investigation of rostral MFC, our finding of reduced GABA in the rostral MFC was suggestive of a GABA deficit, although the failure to survive correction for multiple comparisons suggests caution around any definitive conclusions.

### Limitations

The present study was not without limitations. Due to the limited number of studies available, particularly of bipolar disorder and UHR individuals, we were unable to examine GABA+ concentrations in some regions of the MFC, in some of our clinical populations of interest. Additionally, many studies that were included in our meta-analyses had small sample sizes, and were therefore likely low or underpowered, which can lead to inflated effect sizes and reduced the likelihood that significant results are true effects(95). While we found no evidence of publication bias, i.e., the tendency for studies with positive findings to be more likely to be published, however it is possible that the pooled effects we obtained from our meta-analyses are similarly inflated.

In the current study, classification of voxels into the appropriate subregions was performed by one of the authors, who visually examined the voxel location figures included in the articles or classified according to written descriptions if figures were not included. Classifications were then reviewed and discussed with the remaining authors, and re-classified if necessary. Despite our best efforts, the classifications were somewhat subjective, and accuracy depended on the example image, or description provided in the original article. Indeed, as noted above, when we compared our classification of MFC voxels with those of Nakahara et al.(19), and Kumar et al.(18), we found several differences. These differences undoubtedly impact the results.

In-line with many other meta-analyses of GABA concentrations in psychosis spectrum disorders and depression, we found evidence for moderate to large amounts of between-study heterogeneity. This heterogeneity likely reflects several factors, such as differences in patient sample characteristics, as well as differences in the methodological characteristics of 1H-MRS data collection. We set out to reduce heterogeneity by pooling effects from overlapping voxels placed in small, more functionally homogeneous regions and performing subgroup analyses based on illness duration in psychosis and current symptom profile in depression. While the result yielded voxel clusters with reduced heterogeneity and significant differences, such as the mid- and posterior-MFC in psychosis, other voxels, such as the rostral-MFC in psychosis, still exhibited high heterogeneity.

Finally, a limitation of all 1H-MRS studies is that while the technique allows the non-invasive, in vivo quantification of GABA, it is not able to distinguish between intra- and extra-cellular GABA pools, making interpretation and reconciliation with post-mortem findings difficult.

## Conclusion

The present study utilizes a series of 1H-MRS meta-analyses to reveal medial frontal GABA+ alterations in psychosis spectrum disorders and clarifies prior inconsistent findings. While more studies are required to fully explore the sub-regions of the MFC in bipolar disorder and in individuals at risk for developing psychosis, these results suggest abnormal GABAergic transmission in psychosis, and support the role of the GABA system as a target for therapeutics.

## Data Availability

All data produced on the present work are contained in the manuscript

## ACKNOWLEDGEMENTS

SFT is supported by R01MH118634-01 from NIMH

## REFERENCES

1. Lewis DA, Hashimoto T, Volk DW (2005, April): Cortical inhibitory neurons and schizophrenia. Nature Reviews Neuroscience, vol. 6. Nature Publishing Group, pp 312–324.

2. Hashimoto T, Volk DW, Eggan SM, Mirnics K, Pierri JN, Sun Z, et al. (2003): Gene expression deficits in a subclass of GABA neurons in the prefrontal cortex of subjects with schizophrenia. J Neurosci 23: 6315–6326.

3. Volk DW, Austin MC, Pierri JN, Sampson AR, Lewis DA (2000): Decreased glutamic acid decarboxylase67 messenger RNA expression in a subset of prefrontal cortical γ-aminobutyric acid neurons in subjects with schizophrenia. Arch Gen Psychiatry 57: 237–245.

4. Akbarian S, Kim JJ, Potkin SG, Hagman JO, Tafazzoli A, Bunney WE, Jones EG (1995): Gene Expression for Glutamic Acid Decarboxylase is Reduced without Loss of Neurons in Prefrontal Cortex of Schizophrenics. Arch Gen Psychiatry 52: 258–266.

5. Petty F (1994): Plasma concentrations of γ-aminobutyric acid (GABA) and mood disorders: A blood test for manic depressive disease? Clinical Chemistry, vol. 40 40: 296–302.

6. Petty F, Kramer GL, Fulton M, Davis L, Rush AJ (1995): Stability of plasma GABA at four-year follow-up in patients with primary unipolar depression. Biol Psychiatry 37: 806–810.

7. Gold BI, Bowers MB, Roth RH, Sweeney DW (1980): GABA levels in CSF of patients with psychiatric disorders. Am J Psychiatry 137: 362–364.

8. Gerner RH, Hare TA (1981): CSF GABA in normal subjects and patients with depression, schizophrenia, mania, and anorexia nervosa. Am J Psychiatry 138: 1098–1101.

9. Kasa K, Otsuki S, Yamamoto M, Sato M, Kuroda H, Ogawa N (1982): Cerebrospinal fluid γ-aminobutyric acid and homovanillic acid in depressive disorders. Biol Psychiatry 17: 877–883.

10. Fee C, Banasr M, Sibille E (2017, October 15): Somatostatin-Positive Gamma-Aminobutyric Acid Interneuron Deficits in Depression: Cortical Microcircuit and Therapeutic Perspectives. Biological Psychiatry, vol. 82. Elsevier, pp 549–559.

11. Mescher M, Tannus A, O’Neil Johnson M, Garwood M (1996): Solvent suppression using selective echo dephasing. J Magn Reson - Ser A 123: 226–229.

12. Mescher M, Merkle H, Kirsch J, Garwood M, Gruetter R (1998): Simultaneous in vivo spectral editing and water suppression. NMR Biomed 11: 266–272.

13. Öngür D, Prescot AP, McCarthy J, Cohen BM, Renshaw PF (2010): Elevated gamma-aminobutyric acid levels in chronic schizophrenia. Biol Psychiatry 68: 667–670.

14. Kegeles LS, Mao X, Stanford AD, Girgis R, Ojeil N, Xu X, et al. (2012): Elevated prefrontal cortex γ-aminobutyric acid and glutamate-glutamine levels in schizophrenia measured in vivo with proton magnetic resonance spectroscopy. Arch Gen Psychiatry 69: 449–459.

15. Rowland LM, Kontson K, West J, Edden RA, Zhu H, Wijtenburg SA, et al. (2013): In vivo measurements of glutamate, GABA, and NAAG in schizophrenia. Schizophr Bull 39: 1096–1104.

16. Kelemen O, Kiss I, Benedek G, Kéri S (2013): Perceptual and cognitive effects of antipsychotics in first-episode schizophrenia: The potential impact of GABA concentration in the visual cortex. Prog Neuro-Psychopharmacology Biol Psychiatry 47: 13–19.

17. Tayoshi S, Nakataki M, Sumitani S, Taniguchi K, Shibuya-Tayoshi S, Numata S, et al. (2010): GABA concentration in schizophrenia patients and the effects of antipsychotic medication: A proton magnetic resonance spectroscopy study. Schizophr Res 117: 83–91.

18. Kumar V, Vajawat B, Rao NP (2021): Frontal GABA in schizophrenia: A meta-analysis of 1H-MRS studies. World J Biol Psychiatry 22: 1–13.

19. Nakahara T, Tsugawa S, Noda Y, Ueno F, Honda S, Kinjo M, et al. (2021): Glutamatergic and GABAergic metabolite levels in schizophrenia-spectrum disorders: a meta-analysis of 1H-magnetic resonance spectroscopy studies. Molecular Psychiatry. https://doi.org/10.1038/s41380-021-01297-6

20. Schür RR, Draisma LW, Wijnen JP, Boks MP, Koevoets MGJC, Joëls M, et al. (2016, September 1): Brain GABA levels across psychiatric disorders: A systematic literature review and meta-analysis of 1H-MRS studies. Human Brain Mapping, vol. 37. John Wiley and Sons Inc., pp 3337–3352.

21. Sydnor VJ, Roalf DR (2020): A meta-analysis of ultra-high field glutamate, glutamine, GABA and glutathione 1HMRS in psychosis: Implications for studies of psychosis risk. Schizophr Res 226: 61–69.

22. Egerton A, Modinos G, Ferrera D, McGuire P (2017): Neuroimaging studies of GABA in schizophrenia: A systematic review with meta-analysis. Translational Psychiatry, vol. 7. https://doi.org/10.1038/tp.2017.124

23. Boy F, Evans CJ, Edden RAE, Singh KD, Husain M, Sumner P (2010): Individual differences in subconscious motor control predicted by GABA concentration in SMA. Curr Biol 20: 1779–1785.

24. Grachev ID, Vania Apkarian A (2001): Aging alters regional multichemical profile of the human brain: An in vivo 1H-MRS study of young versus middle-aged subjects. J Neurochem 76: 582–593.

25. Greenhouse I, Noah S, Maddock RJ, Ivry RB (2016): Individual differences in GABA content are reliable but are not uniform across the human cortex. Neuroimage 139: 1–7.

26. Vega A de la, Chang LJ, Banich MT, Wager TD, Yarkoni T (2016): Large-Scale Meta-Analysis of Human Medial Frontal Cortex Reveals Tripartite Functional Organization. J Neurosci 36: 6553–6562.

27. Taylor SF, Tso IF (2015): GABA abnormalities in schizophrenia: A methodological review of in vivo studies. Schizophrenia Research, vol. 167. pp 84–90.

28. Romeo B, Choucha W, Fossati P, Rotge JY (2018): Meta-analysis of central and peripheral γ-aminobutyric acid levels in patients with unipolar and bipolar depression. J Psychiatry Neurosci 43: 58–66.

29. Scotti-Muzzi E, Umla-Runge K, Soeiro-de-Souza MG (2021): Anterior cingulate cortex neurometabolites in bipolar disorder are influenced by mood state and medication: A meta-analysis of 1H-MRS studies. European Neuropsychopharmacology, vol. 47. pp 62–73.

30. Godfrey KEM, Gardner AC, Kwon S, Chea W, Muthukumaraswamy SD (2018): Differences in excitatory and inhibitory neurotransmitter levels between depressed patients and healthy controls: A systematic review and meta-analysis. Journal of Psychiatric Research, vol. 105. pp 33–44.

31. Wenneberg C, Glenthøj BY, Hjorthøj C, Buchardt Zingenberg FJ, Glenthøj LB, Rostrup E, et al. (2020): Cerebral glutamate and GABA levels in high-risk of psychosis states: A focused review and meta-analysis of 1H-MRS studies. Schizophrenia Research, vol. 215. pp 38–48.

32. Conn VS, Valentine JC, Cooper HM, Rantz MJ, Vs C, Jc V, et al. (2003): Grey literature in meta-analyses. Nursing Research, vol. 52. Nurs Res, pp 256–261.

33. Viechtbauer W (2010): Conducting meta-analyses in R with the metafor. J Stat Softw 36: 1–48.

34. Huedo-Medina TB, Sánchez-Meca J, Marín-Martínez F, Botella J (2006): Assessing heterogeneity in meta-analysis: Q statistic or I 2 Index? Psychol Methods 11: 193–206.

35. Eggers HC, Lipa P, Buschbeck B (1997): Sensitive test for models of bose-einstein correlations. Phys Rev Lett 79: 197–200.

36. Xia M, Wang J, Sheng J, Tang Y, Li C, Lim K, et al. (2018): Effect of Electroconvulsive Therapy on Medial Prefrontal γ-Aminobutyric Acid among Schizophrenia Patients: A Proton Magnetic Resonance Spectroscopy Study. J ECT 34: 227–232.

37. Yang Z, Zhu Y, Song Z, Mei L, Zhang J, Chen T, et al. (2015): Comparison of the density of gamma-aminobutyric acid in the ventromedial prefrontal cortex of patients with first-episode psychosis and healthy controls. Shanghai Arch Psychiatry 27: 341–347.

38. Marsman A, Mandl RCW, Klomp DWJ, Bohlken MM, Boer VO, Andreychenko A, et al. (2014): GABA and glutamate in schizophrenia: A 7 T 1H-MRS study. NeuroImage Clin 6: 398–407.

39. Bojesen KB, Ebdrup BH, Jessen K, Sigvard A, Tangmose K, Edden RAE, et al. (2020): Treatment response after 6 and 26 weeks is related to baseline glutamate and GABA levels in antipsychotic-naïve patients with psychosis. Psychol Med 50: 2182–2193.

40. Brandt AS, Unschuld PG, Pradhan S, Lim IAL, Churchill G, Harris AD, et al. (2016): Age-related changes in anterior cingulate cortex glutamate in schizophrenia: A 1H MRS Study at 7 Tesla. Schizophr Res 172: 101–105.

41. Goto N, Yoshimura R, Moriya J, Kakeda S, Ueda N, Ikenouchi-Sugita A, et al. (2009): Reduction of brain γ-aminobutyric acid (GABA) concentrations in early-stage schizophrenia patients: 3T Proton MRS study. Schizophrenia Research, vol. 112. pp 192–193.

42. Reid MA, Salibi N, White DM, Gawne TJ, Denney TS, Lahti AC (2019): 7T Proton Magnetic Resonance Spectroscopy of the Anterior Cingulate Cortex in First-Episode Schizophrenia. Schizophr Bull 45: 180–189.

43. Rowland LM, Krause BW, Wijtenburg SA, McMahon RP, Chiappelli J, Nugent KL, et al. (2016): Medial frontal GABA is lower in older schizophrenia: A MEGA-PRESS with macromolecule suppression study. Mol Psychiatry 21: 198–204.

44. Wijtenburg SA, Wang M, Korenic SA, Chen S, Barker PB, Rowland LM (2021): Metabolite Alterations in Adults With Schizophrenia, First Degree Relatives, and Healthy Controls: A Multi-Region 7T MRS Study. Front Psychiatry 12. https://doi.org/10.3389/fpsyt.2021.656459

45. Hjelmervik H, Craven AR, Sinceviciute I, Johnsen E, Kompus K, Bless JJ, et al. (2020): Intra-Regional Glu-GABA vs Inter-Regional Glu-Glu Imbalance: A 1H-MRS Study of the Neurochemistry of Auditory Verbal Hallucinations in Schizophrenia. Schizophr Bull 46: 633–642.

46. Marenco S, Meyer C, Kuo S, Van Der Veen JW, Shen J, DeJong K, et al. (2016): Prefrontal GABA levels measured with magnetic resonance spectroscopy in patients with psychosis and unaffected siblings. American Journal of Psychiatry, vol. 173 173: 527–534.

47. Wang JJ, Wang JJ, Tang Y, Zhang T, Cui H, Xu L, et al. (2016): Reduced γ-Aminobutyric Acid and Glutamate+Glutamine Levels in Drug-Naïve Patients with First-Episode Schizophrenia but Not in Those at Ultrahigh Risk. Neural Plast 2016. https://doi.org/10.1155/2016/3915703

48. Cen H, Xu J, Yang Z, Mei L, Chen T, Zhuo K, et al. (2020): Neurochemical and brain functional changes in the ventromedial prefrontal cortex of first-episode psychosis patients: A combined functional magnetic resonance imaging—proton magnetic resonance spectroscopy study. Aust N Z J Psychiatry 54: 519–527.

49. Chen T, Wang Y, Zhang J, Wang Z, Xu J, Li Y, et al. (2017): Abnormal Concentration of GABA and Glutamate in The Prefrontal Cortex in Schizophrenia.-An in Vivo 1H-MRS Study. Shanghai Arch Psychiatry 29: 277–286.

50. Chiu PW, Lui SSY, Hung KSY, Chan RCK, Chan Q, Sham PC, et al. (2018): In vivo gamma-aminobutyric acid and glutamate levels in people with first-episode schizophrenia: A proton magnetic resonance spectroscopy study. Schizophr Res 193: 295–303.

51. de la Fuente-Sandoval C, Reyes-Madrigal F, Mao X, León-Ortiz P, Rodríguez-Mayoral O, Jung-Cook H, et al. (2018): Prefrontal and Striatal Gamma-Aminobutyric Acid Levels and the Effect of Antipsychotic Treatment in First-Episode Psychosis Patients. Biol Psychiatry 83: 475–483.

52. Ragland JD, Maddock RJ, Hurtado MY, Tanase C, Lesh TA, Niendam TA, et al. (2020): Disrupted GABAergic facilitation of working memory performance in people with schizophrenia. NeuroImage Clin 25. https://doi.org/10.1016/j.nicl.2019.102127

53. Simmonite M, Taylor SF (2021): GABA concentrations in first episode psychosis and attenuated psychosis syndrome. Manuscript in Preparation.

54. De La Fuente-Sandoval C, Reyes-Madrigal F, Mao X, León-Ortiz P, Rodríguez-Mayoral O, Solís-Vivanco R, et al. (2015): Cortico-striatal GABAergic and glutamatergic dysregulations in subjects at ultra-high risk for psychosis investigated with proton magnetic resonance spectroscopy. Int J Neuropsychopharmacol 19: 1–10.

55. Menschikov PE, Semenova NA, Ublinskiy M V., Akhadov TA, Keshishyan RA, Lebedeva IS, et al. (2016): 1H-MRS and MEGA-PRESS pulse sequence in the study of balance of inhibitory and excitatory neurotransmitters in the human brain of ultra-high risk of schizophrenia patients. Dokl Biochem Biophys 468: 168–172.

56. Da Silva T, Hafizi S, Rusjan PM, Houle S, Wilson AA, Prce I, et al. (2019): GABA levels and TSPO expression in people at clinical high risk for psychosis and healthy volunteers: A PET-MRS study. J Psychiatry Neurosci 44: 111–119.

57. Modinos G, Simsek F, Azis M, Bossong M, Bonoldi I, Samson C, et al. (2018): Prefrontal GABA levels, hippocampal resting perfusion and the risk of psychosis. Neuropsychopharmacology 43: 2652–2659.

58. Wenneberg C, Glenthøj BY, Glenthøj LB, Fagerlund B, Krakauer K, Kristensen TD, et al. (2020): Baseline measures of cerebral glutamate and GABA levels in individuals at ultrahigh risk for psychosis: Implications for clinical outcome after 12 months. Eur Psychiatry 63. https://doi.org/10.1192/j.eurpsy.2020.77

59. Brennan BP, Admon R, Perriello C, LaFlamme EM, Athey AJ, Pizzagalli DA, et al. (2017): Acute change in anterior cingulate cortex GABA, but not glutamine/glutamate, mediates antidepressant response to citalopram. Psychiatry Res - Neuroimaging 269: 9–16.

60. Gabbay V, Mao X, Klein RG, Ely BA, Babb JS, Panzer AM, et al. (2012): Anterior cingulate cortex γ-aminobutyric acid in depressed adolescents: Relationship to anhedonia. Arch Gen Psychiatry 69: 139–149.

61. Deligiannidis KM, Fales CL, Kroll-Desrosiers AR, Shaffer SA, Villamarin V, Tan Y, et al. (2019): Resting-state functional connectivity, cortical GABA, and neuroactive steroids in peripartum and peripartum depressed women: a functional magnetic resonance imaging and spectroscopy study. Neuropsychopharmacology 44: 546–554.

62. Draganov M, Vives-Gilabert Y, de Diego-Adeliño J, Vicent-Gil M, Puigdemont D, Portella MJ (2020): Glutamatergic and GABA-ergic abnormalities in First-episode depression. A 1-year follow-up 1H-MR spectroscopic study. J Affect Disord 266: 572–577.

63. Knudsen MK, Near J, Blicher AB, Videbech P, Blicher JU (2019): Magnetic resonance (MR) spectroscopic measurement of γ-aminobutyric acid (GABA) in major depression before and after electroconvulsive therapy. Acta Neuropsychiatr 31: 17–26.

64. Baeken C, Lefaucheur JP, Van Schuerbeek P (2017): The impact of accelerated high frequency rTMS on brain neurochemicals in treatment-resistant depression: Insights from 1H MR spectroscopy. Clin Neurophysiol 128: 1664–1672.

65. Bhagwagar Z, Wylezinska M, Jezzard P, Evans J, Boorman E, Matthews PM, Cowen PJ (2008): Low GABA concentrations in occipital cortex and anterior cingulate cortex in medication-free, recovered depressed patients. Int J Neuropsychopharmacol 11: 255–260.

66. Persson J, Wall A, Weis J, Gingnell M, Antoni G, Lubberink M, Bodén R (2021): Inhibitory and excitatory neurotransmitter systems in depressed and healthy: A positron emission tomography and magnetic resonance spectroscopy study. Psychiatry Res - Neuroimaging 315. https://doi.org/10.1016/j.pscychresns.2021.111327

67. Smith GS, Oeltzschner G, Gould NF, Leoutsakos JMS, Nassery N, Joo JH, et al. (2021): Neurotransmitters and Neurometabolites in Late-Life Depression: A Preliminary Magnetic Resonance Spectroscopy Study at 7T. J Affect Disord 279: 417–425.

68. Benson KL, Bottary R, Schoerning L, Baer L, Gonenc A, Eric Jensen J, Winkelman JW (2020): 1H MRS Measurement of Cortical GABA and Glutamate in Primary Insomnia and Major Depressive Disorder: Relationship to Sleep Quality and Depression Severity. J Affect Disord 274: 624–631.

69. Hasler G, Neumeister A, Van Der Veen JW, Tumonis T, Bain EE, Shen J, et al. (2005): Normal prefrontal gamma-aminobutyric acid levels in remitted depressed subjects determined by proton magnetic resonance spectroscopy. Biol Psychiatry 58: 969–973.

70. Hasler G, Van Der Veen JW, Tumonis T, Meyers N, Shen J, Drevets WC (2007): Reduced prefrontal glutamate/glutamine and γ-aminobutyric acid levels in major depression determined using proton magnetic resonance spectroscopy. Arch Gen Psychiatry 64: 193–200.

71. Ironside M, Moser AD, Holsen LM, Zuo CS, D. F, Perlo S, et al. (2021): Reductions in rostral anterior cingulate GABA are associated with stress circuitry in females with major depression: a multimodal imaging investigation. Neuropsychopharmacology 46: 2188–2196.

72. Kantrowitz JT, Dong Z, Milak MS, Rashid R, Kegeles LS, Javitt DC, et al. (2021): Ventromedial prefrontal cortex/anterior cingulate cortex Glx, glutamate, and GABA levels in medication-free major depressive disorder. Transl Psychiatry 11. https://doi.org/10.1038/s41398-021-01541-1

73. Price RB, Shungu DC, Mao X, Nestadt P, Kelly C, Collins KA, et al. (2009): Amino Acid Neurotransmitters Assessed by Proton Magnetic Resonance Spectroscopy: Relationship to Treatment Resistance in Major Depressive Disorder. Biol Psychiatry 65: 792–800.

74. Walter M, Henning A, Grimm S, Schulte RF, Beck J, Dydak U, et al. (2009): The relationship between aberrant neuronal activation in the pregenual anterior cingulate, altered glutamatergic metabolism, and anhedonia in major depression. Arch Gen Psychiatry 66: 478–486.

75. Wang Z, Zhang A, Zhao B, Gan J, Wang G, Gao F, et al. (2016): GABA+ levels in postmenopausal women with mild-to-moderate depression A preliminary study. Med (United States) 95. https://doi.org/10.1097/MD.0000000000004918

76. Zhang Z, Fan Q, Bai Y, Wang Z, Zhang H, Xiao Z (2016): Brain gamma-aminobutyric acid (GABA) concentration of the prefrontal lobe in unmedicated patients with Obsessivecompulsive disorder: a research of magnetic resonance spectroscopy. Shanghai Arch Psychiatry 28: 263–270.

77. Brady RO, Mccarthy JM, Prescot AP, Jensen JE, Cooper AJ, Cohen BM, et al. (2013): Brain gamma-aminobutyric acid (GABA) abnormalities in bipolar disorder. Bipolar Disord 15: 434–439.

78. Godlewska BR, Yip SW, Near J, Goodwin GM, Cowen PJ (2014): Cortical glutathione levels in young people with bipolar disorder: A pilot study using magnetic resonance spectroscopy. Psychopharmacology (Berl) 231: 327–332.

79. Wang PW, Sailasuta N, Chandler RA, Ketter TA (2006): Magnetic resonance spectroscopic measurement of cerebral gamma-aminobutyric acid concentrations in patients with bipolar disorders. Acta Neuropsychiatr 18: 120–126.

80. Huber RS, Kondo DG, Shi XF, Prescot AP, Clark E, Renshaw PF, Yurgelun-Todd DA (2018): Relationship of executive functioning deficits to N-acetyl aspartate (NAA) and gamma-aminobutyric acid (GABA) in youth with bipolar disorder. J Affect Disord 225: 71–78.

81. Prisciandaro JJ, Tolliver BK, Prescot AP, Brenner HM, Renshaw PF, Brown TR, Anton RF (2017): Unique prefrontal GABA and glutamate disturbances in co-occurring bipolar disorder and alcohol dependence. Transl Psychiatry 7. https://doi.org/10.1038/tp.2017.141

82. Soeiro-de-Souza MG, Henning A, Machado-Vieira R, Moreno RA, Pastorello BF, da Costa Leite C, et al. (2015): Anterior cingulate Glutamate-Glutamine cycle metabolites are altered in euthymic bipolar I disorder. Eur Neuropsychopharmacol 25: 2221–2229.

83. Pomarol-Clotet E, Canales-Rodríguez EJ, Salvador R, Sarró S, Gomar JJ, Vila F, et al. (2010): Medial prefrontal cortex pathology in schizophrenia as revealed by convergent findings from multimodal imaging. Mol Psychiatry 15: 823–830.

84. Ridderinkhof KR, Ullsperger M, Crone EA, Nieuwenhuis S (2004, October 15): The role of the medial frontal cortex in cognitive control. Science, vol. 306. Science, pp 443–447.

85. Taylor SF, Liberzon I (2007): Neural correlates of emotion regulation in psychopathology. Trends Cogn Sci 11: 413–418.

86. Waugh C, Lemus M, … IG-C and A, 2014 U (n.d.): The role of the medial frontal cortex in the maintenance of emotional states. academic.oup.com. Retrieved February 1, 2022, from https://academic.oup.com/scan/article-abstract/9/12/2001/1619973

87. Luu P, Flaisch T, Tucker DM (2000): Medial frontal cortex in action monitoring. J Neurosci 20: 464–469.

88. Rushworth MFS, Walton ME, Kennerley SW, Bannerman DM (2004): Action sets and decisions in the medial frontal cortex. Trends in Cognitive Sciences, vol. 8. pp 410–417.

89. Barch DM, Ceaser A (2012, January 1): Cognition in schizophrenia: Core psychological and neural mechanisms. Trends in Cognitive Sciences, vol. 16. Elsevier Current Trends, pp 27–34.

90. Minzenberg MJ, Laird AR, Thelen S, Carter CS, Glahn DC (2009): Meta-analysis of 41 functional neuroimaging studies of executive function in schizophrenia. Arch Gen Psychiatry 66: 811–822.

91. Petty F, Sherman AD (1984): Plasma GABA levels in psychiatric illness. J Affect Disord 6: 131–138.

92. Petty F, Schlesser MA (1981): Plasma GABA in affective illness: A preliminary investigation. J Affect Disord 3: 339–343.

93. Perrine SA, Ghoddoussi F, Michaels MS, Sheikh IS, McKelvey G, Galloway MP (2014): Ketamine reverses stress-induced depression-like behavior and increased GABA levels in the anterior cingulate: An 11.7T 1H-MRS study in rats. Prog Neuro-Psychopharmacology Biol Psychiatry 51: 9–15.

94. Rodriguez CI, Kegeles LS, Levinson A, Ogden RT, Mao X, Milak MS, et al. (2015): In vivo effects of ketamine on glutamate-glutamine and gamma-aminobutyric acid in obsessive-compulsive disorder: Proof of concept. Psychiatry Res - Neuroimaging 233: 141–147.

95. Button KS, Ioannidis JPA, Mokrysz C, Nosek BA, Flint J, Robinson ESJ, Munafò MR (2013): Power failure: Why small sample size undermines the reliability of neuroscience. Nat Rev Neurosci 14: 365–376.

