## Supplemental material for "MEDIAL FRONTAL CORTEX GAMMA-AMINOBUTYRIC ACID CONCENTRATIONS IN PSYCHOSIS AND MOOD DISORDERS: A META-ANALYSIS OF PROTON MAGNETIC RESONANCE SPECTROSCOPY STUDIES"

| Supplementary Table 1: Characteristics of included Psychosis studies | | | | | | | | | | | |
| --- | --- | --- | --- | --- | --- | --- | --- | --- | --- | --- | --- |
| Study | Case | | Control | | Gender (%male) | Currently Medicated | Illness duration | MRS technique | Field Strength | Reference Metabolite | Analysis software |
|  | n | Mean age  ± SD | n | Mean age ± SD |  |  |  |  |  |  |  |
| ***Rostral MFC*** | | | | | | | | | | | |
| Cen et al., 2020 (1) | 23 | 27.0 ± 6.5 | 26 | 25.9 ± 4.6 | 39.1 | Unmedicated | Acute | MEGA-PRESS | 3 | Water | LCModel |
| Chen et al., 2017 (2) | 24 | 28.8 ± 8.3 | 24 | 26.6 ± 4.7 | 41.7 | Unmedicated | Acute | MEGA-PRESS | 3 | Water | LCModel |
| Chiu et al., 2018 (3) | 19 | 29.1 ± 6.7 | 14 | 27.7 ± 5.9 | 57.4 | Medicated | Acute | MEGA-PRESS | 3 | Water | Gannet 2.0 |
| De la Fuente-Sandoval et al., 2018 (4) | 28 | 23.0 ± 6.1 | 18 | 23.0 ± 3.8 | 71.4 | Unmedicated | Acute | J-editing | 3 | Water | Inhouse software |
| Kegeles et al., 2012 (medicated) (5) | 16 | 32.0 ± 10.0 | 11 | 33.0 ± 8.0 | 68.8 | Medicated | Chronic | J-editing | 3 | Water | Inhouse software |
| Kegeles et al., 2012 (unmedicated) (5) | 16 | 32.0 ± 11.0 | 11 | 33.0 ± 8.0 | 68.8 | Unmedicated | Chronic | J-editing | 3 | Water | Inhouse software |
| Ragland et al., 2020 (medicated) (6) | 29 | 23.5 ± 4.6 | 25 | 24.2 ± 4.7 | 70.0 | Medicated | Acute | MEGA-PRESS | 3 | Cr | jMRUI 4.0 |
| Ragland et al., 2020 (unmedicated) (6) | 9 | 23.5 ± 4.6 | 24 | 24.2 ± 4.7 | 70.0 | Unmedicated | Acute | MEGA-PRESS | 3 | Cr | jMRUI 4.0 |
| Simmonite et al., 2022 (7) | 14 | 21.6 ± 2.6 | 15 | 21.6 ± 3.6 | 64.3 | Mixed | Acute | MEGA-PRESS | 3 | Water | Gannet 3.0 |
| Wang et al., 2016 (8) | 16 | 22.1 ± 5.5 | 23 | 22.5 ± 5.5 | 50.0 | Unmedicated | Acute | MEGA-PRESS | 3 | Water | LCModel |
| Xia et al., 2018 (drug group baseline) (9) | 17 | 31.0 ± 7.7 | 10 | 30.9 ± 5.3 | 47.0 | Medicated | Chronic | MEGA-PRESS | 3 | Cr | Gannet 2.0 |
| Xia et al., 2018 (ECT group baseline) (9) | 14 | 27.6 ± 7.3 | 9 | 30.9 ± 5.3 | 50.0 | Medicated | Chronic | MEGA-PRESS | 3 | Cr | Gannet 2.0 |
| Yang et al., 2015 (10) | 22 | 26.1 ± 5.8 | 23 | 25.5 ± 4.4 | 40.9 | Unmedicated | Acute | MEGA-PRESS | 3 | Water | LCModel |
| ***Rostral-Mid MFC*** | | | | | | | | | | | |
| Marsman et al., 2014 (11) | 13 | 27.6 ± 6.1 | 19 | 27.7 ± 5.3 | 76.5 | Medicated | Chronic | MEGA-sLASER | 7 | Cr | Inhouse software |
| Rowland et al., 2013 (older) (12) | 10 | 51.1 ± 4.0 | 10 | 49.4 ± 3.9 | 70.0 | Medicated | Chronic | MEGA-PRESS | 3 | Water | csx3 |
| Rowland et al., 2013 (young) (12) | 11 | 30.2 ± 6.6 | 19 | 33.4 ± 6.5 | 81.8 | Medicated | Chronic | MEGA-PRESS | 3 | water | csx3 |
| ***Mid MFC*** |  |  |  |  |  |  |  |  |  |  |  |
| Bojesen et al., 2021 (13) | 37 | 22.7 ± 5.0 | 47 | 22.2 ± 4.1 | 42.9 | Unmedicated | Acute | MEGA-PRESS | 3 | Water | Gannet |
| Brandt et al., 2016 (14) | 24 | 37.5 ± 26.7 | 24 | 36.6 ± 14.5 | 79.2 | Medicated | Chronic | STEAM | 7 | Water | LCModel |
| Goto et al., 2009 (15) | 18 | 29.0 ± 11.0 | 18 | 30.0 ± 11.0 | 50.0 | Medicated | Acute | MEGA-PRESS | 3 | Cr | LCModel |
| Ongur et al., 2010 (16) | 21 | 39.0 ± 10.8 | 19 | 36.3 ± 9.8 | 66.7 | Medicated | Chronic | MEGA-PRESS | 4 | Cr | LCModel |
| Reid et al., 2019 (17) | 21 | 23.3 ± 4.4 | 21 | 23.5 ± 4.4 | 76.2 | Medicated | Acute | STEAM | 7 | Water | LCModel |
| Rowland et al., 2016 (older) (18) | 31 | 48.3 ± 5.8 | 37 | 52.0 ± 6.0 | 61.3 | Mixed | Chronic | MEGA-PRESS | 3 | Water | Gannet 2.0 |
| Rowland et al., 2016 (young) (18) | 29 | 25.7 ± 4.3 | 40 | 25.3 ± 4.6 | 69.0 | Mixed | Chronic | MEGA-PRESS | 3 | Water | Gannet 2.0 |
| Wang et al., 2019 (19) | 81 | 22.3 ± 4.4 | 91 | 25.3 ± 3.9 | 70.4 | Medicated | Acute | STEAM | 7 | Water | LCModel |
| Wijtenburg et al., 2021 (acute) (20) | 18 | 24.3 ± 3.9 | 19 | NR | 68.4 | Medicated | Acute | STEAM | 7 | Water | LCModel |
| Wijtenburg et al., 2021 (chronic) (20) | 21 | 43.1 ± 11.0 | 17 | NR | 47.6 | Medicated | Chronic | STEAM | 7 | Water | LCModel |
| ***Posterior MFC*** | | | | | | | | | | | |
| Hjelmervik et al., 2020 (21) | 37 | 29.8 ± 11.5 | 38 | 30.2 ± 10.2 | NR | Mixed | Chronic | MEGA-PRESS | 3 | Water | LCModel |
| Marenco et al., 2016 (medicated) (22) | 70 | 31.2 ± 9.2 | 92 | 30.6 ± 9.2 | 71.4 | Medicated | Chronic | J-editing | 3 | Cr | Inhouse software |
| Marenco et al., 2016 (unmedicated) (22) | 25 | 28.4 ± 8.7 | 92 | 30.6 ± 9.2 | 72.0 | Unmedicated | Chronic | J-editing | 3 | Cr | Inhouse software |
| Tayoshi et al., 2010 (23) | 38 | 34.0 ± 10.0 | 29 | 34.0 ± 10.2 | 52.6 | Medicated | Chronic | MEGA-PRESS | 3 | Water | LCModel |
| Abbreviations: Cr, creatine; ECT, electroconvulsive therapy; MEGA-PRESS, Mescher-Garwood point resolved spectroscopy; MEGA-sLASER, Mescher-Garwood-semi-localized by adiabatic selective refocusing; MFC, medial frontal cortex; PRESS, point resolved spectroscopy; SD, standard deviation; STEAM, stimulated echo acquisition mode; NR, not reported; | | | | | | | | | | | |

| Supplementary Table 2: Characteristics of included Depression studies | | | | | | | | | | |
| --- | --- | --- | --- | --- | --- | --- | --- | --- | --- | --- |
| Study | Case | | Control | | Gender (%male) | Currently Medicated | MRS technique | Field Strength | Reference Metabolite | Analysis software |
|  | n | Mean age ± SD | n | Mean age ± SD |  |  |  |  |  |  |
| ***Rostral MFC*** |  |  |  |  |  |  |  |  |  |  |
| Brennan et al., 2017 (24) | 17 | 38.5 ± 12.2 | 9 | 38.4 ± 14.1 | 57.9 | Unmedicated | MEGA-PRESS | 3 | Cr | LCModel |
| Gabbay et al., 2012 (anhedonic) (25) | 10 | 18 ± 1.9 | 10 | 16.2 ± 1.6 | 40.0 | Mixed | J-editing | 3 | Water | Inhouse software |
| Gabbay et al., 2012 (non-anhedonic) (25) | 10 | 15.3 ± 2.7 | 11 | 16.2 ± 1.6 | 40.0 | Mixed | J-editing | 3 | Water | Inhouse software |
| Gabbay et al., 2017 (26) | 24 | 16.1 ± 2.6 | 15 | 15.3 ± 2.7 | 54.2 | Unmedicated | J-editing | 3 | Water | Inhouse software |
| Hasler et al., 2005 (27) | 16 | 41 ± 11.6 | 15 | 41.7 ± 12.4 | 33.3 | Unmedicated | J-editing | 3 | Water | MRUI |
| Hasler et al., 2007 (28) | 20 | 34 ± 11.2 | 20 | 34.8 ± 12.4 | 35.0 | Unmedicated | J-editing | 3 | Water | Inhouse software |
| Ironside et al., 2021 (depressed) (29) | 17 | 21.1 ± 1.8 | 7 | 21.5 ± 2.5 | 0.0 | Unmedicated | MEGA-PRESS | 3 | Water | LCModel |
| Ironside et al., 2021 (remitted) (29) | 13 | 21.1 ± 2.12 | 6 | 21.5 ± 2.5 | 0.0 | Unmedicated | MEGA-PRESS | 3 | Water | LCModel |
| Kantrowitz et al., 2021 (female) (30) | 12 | 35 ± 10.4 | 19 | 36.9 ± 9.7 | 0.0 | Unmedicated | PROBE-P/PROBE-J | 3 | NAA | Gannet |
| Kantrowitz et al., 2021 (male) (30) | 22 | 38.4 ± 11 | 13 | 30.7 ± 5.1 | 100.0 | Unmedicated | PROBE-P/PROBE-J | 3 | NAA | Gannet |
| Price et al., 2009 (treatment resistant) (31) | 12 | 46.8 ± 11.8 | 10 | 37.3 ± 13.5 | 56.3 | Unmedicated | J-editing | 3 | Water | Inhouse software |
| Price et al., 2009 (treatment responsive) (31) | 16 | 38.3 ± 12.3 | 11 | 37.3 ± 13.5 | 66.7 | Unmedicated | J-editing | 3 | Water | Inhouse software |
| Walter et al., 2010 (anhedonic) (32) | 4 | 40 ± NR | 6 | 34.6 ± NR | 42.1 | Unmedicated | J-PRESS | 3 | Cr | ProFit |
| Walter et al., 2010 (non-anhedonic) (32) | 5 | 40 ± NR | 7 | 34.6 ± NR | 42.1 | Unmedicated | J-PRESS | 3 | Cr | ProFit |
| Wang et al., 2019 (33) | 19 | 47.7 ± 1.9 | 71 | 46.8 ± 2.0 | 0.0 | NR | MEGA-PRESS | 3 | Water | TARQUIN |
| Zhang et al., 2016 (34) | 11 | 34.1 ± 8.8 | 11 | 33.6 ±7.2 | 0.0 | Mixed | MEGA-PRESS | 3 | Water | LCModel |
| ***Rostral-Mid MFC*** |  |  |  |  |  |  |  |  |  |  |
| Deligiannidis et al., 2019 (35) | 25 | 28.6 ± 4.9 | 28 | 29.0 ± 5.0 | 0 | Unmedicated | MEGA-PRESS | 3 | Cr | Gannet |
| Draganov et al., 2020 (36) | 23 | 37.3 ± 10.8 | 54 | 41.8 ± 10.1 | 41.9 | Unmedicated | PRESS | 3 | Water | TARQUIN |
| Hasler et al., 2005 (27) | 16 | 41.0 ± 11.6 | 15 | 41.7 ± 12.4 | 33.3 | Unmedicated | J-editing | 3 | Water | MRUI |
| Hasler et al., 2007 (28) | 20 | 34.0 ± 11.2 | 20 | 34.8 ± 12.4 | 35.0 | Unmedicated | J-editing | 3 | Water | Inhouse software |
| Knudsen et al., 2019 (37) | 10 | 38.4 ± 10.9 | 10 | 38.8 ± 10.8 | 40.0 | Medicated | SPECIAL | 3 | Cr | LCModel |
| ***Mid MFC*** |  |  |  |  |  |  |  |  |  |  |
| Baeken et al., 2017 (38) | 18 | 47.2 ± 12.5 | 18 | 45.8 ± 12.3 | 33.3 | Mixed | PRESS | 3 | Water | jMRUI 5.1 |
| Benson et al., 2020 (39) | 41 | 33.2 ± 14.4 | 20 | 33.9 ± 14.6 | 19.5 | Unmedicated | MEGA-PRESS | 4 | Cr | LCModel |
| Bhagwagar et al., 2008 (40) | 12 | 40.6 ± 4.2 | 11 | 34.3 ± 4.1 | 33.3 | Unmedicated | MEGA-PRESS | 3 | Cr | LCModel |
| Persson et al., 2021 (41) | 42 | 29.2 ± 9.4 | 45 | 29.5 ± 11.2 | 50.0 | Mixed | MEGA-PRESS | 3 | Cr | Gannet 3.0 |
| Smith et al., 2021 (42) | 9 | 70.0 ± 7.0 | 9 | 67.0 ± 7.0 | 44.4 | Medicated | STEAM | 7 | Cr | LCModel |
| Wang et al., 2016 (43) | 19 | 53.9 ± 2.6 | 13 | 52.6 ± 2.18 | 0 | Unmedicated | MEGA-PRESS | 3 | Water | Gannet |
| Abbreviations: Cr, creatine; MEGA-PRESS, Mescher-Garwood point resolved spectroscopy; MFC, medial frontal cortex; PRESS, point resolved spectroscopy; SD, standard deviation; SPECIAL, spin echo full intensity-acquired localized spectroscopy; STEAM, stimulated echo acquisition mode; NAA, N-acetyl aspartate; NR, not reported. | | | | | | | | | | |

| Supplementary Table 3: Characteristics of included Bipolar disorder studies | | | | | | | | | | |
| --- | --- | --- | --- | --- | --- | --- | --- | --- | --- | --- |
| Study | Case | | Control | | Gender (%male) | Currently Medicated | MRS technique | Field Strength | Reference Metabolite | Analysis software |
|  | n | Mean age ± SD | n | Mean age ± SD |  |  |  |  |  |  |
| ***Rostral MFC*** |  |  |  |  |  |  |  |  |  |  |
| Brady et al., 2013 (medicated) (44) | 10 | 32.6 ± 13.6 | 7 | 36.9 ± 10.4 | 57.1 | Medicated | MEGA-PRESS | 4 | Cr | LCModel |
| Brady et al., 2013 (unmedicated) (44) | 4 | 32.6 ± 13.6 | 7 | 36.9 ± 10.4 | 57.1 | Unmedicated | MEGA-PRESS | 4 | Cr | LCModel |
| Godlewska et al., 2014 (45) | 13 | 23.8 ± 3.6 | 11 | 21.9 ± 2.7 | 46.2 | Unmedicated | SPECIAL | 3 | Cr | LCModel 6.2-2B |
| Wang et al., 2006 (46) | 15 | 35.7 ± 11.4 | 6 | 41.7 ± 21.2 | 80.0 | Mixed | J-editing | 3 | Cr | SAGE |
| ***Mid MFC*** |  |  |  |  |  |  |  |  |  |  |
| Huber et al., 2018 (47) | 19 | 17.5 ± 2.6 | 10 | 19.0 ± 1.6 | 31.6 | NR | J-PRESS | 3 | Water | ProFit |
| Priscandaro et al., 2017 (48) | 18 | 36.3 ± 11.4 | 15 | 38.0 ± 11.1 | 55.0 | Mixed | J-RES | 3 | Water | ProFit |
| Soeiro-de-Souza et al., 2015 (49) | 50 | 31.7 ± 9.1 | 38 | 25.7 ± 5.7 | 38.0 | Mixed | J-PRESS | 3 | Cr | ProFit |
| Abbreviations: Cr, creatine; MEGA-PRESS, Meschler-Garwood point resolved spectroscopy; MFC, medial frontal cortex; PRESS, point resolved spectroscopy; SD, standard deviation; SPECIAL, spin echo full intensity-acquired localized spectroscopy; NR, not reported | | | | | | | | | | |

| Supplementary Table 4: Characteristics of included Ultra High-Risk studies | | | | | | | | | | |
| --- | --- | --- | --- | --- | --- | --- | --- | --- | --- | --- |
| Study | Case | | Control | | Gender (%male) | Currently Medicated | MRS technique | Field Strength | Reference Metabolite | Analysis software |
|  | n | Mean age ± SD | n | Mean age ± SD |  |  |  |  |  |  |
| ***Rostral MFC*** |  |  |  |  |  |  |  |  |  |  |
| De la Fuente-Sandoval et al., 2015 (50) | 23 | 20.7 ± 4.1 | 24 | 21.4 ± 3.3 | 65.22 | Unmedicated | J-editing | 3 | Water | Inhouse software |
| Menschikov et al., 2016 (51) | 21 | NR | 26 | NR | 100.0 | NR | MEGA-PRESS | 3 | Cr | jMRUI 5.1a |
| Simmonite et al., 2022 (7) | 7 | 19.9 ± 3.6 | 15 | 21.6 ± 21.6 | 71.43 | Mixed | MEGA-PRESS | 3 | Water | Gannet 3.1 |
| Wang et al., 2016 (8) | 21 | 21.1 ± 5.7 | 23 | 22.5 ± 22.5 | 57.14 | Unmedicated | MEGA-PRESS | 3 | Water | LCModel |
| ***Mid MFC*** |  |  |  |  |  |  |  |  |  |  |
| Wenneberg et al., 2020 (medicated) (52) | 54 | 23.9 ± 4.2 | 26 | 25.3 ± 5.2 | N.R. | Medicated | MEGA-PRESS | 3 | Water | Gannet 3.0 |
| Wenneberg et al., 2020 (unmedicated) (52) | 47 | 23.9 ± 4.2 | 27 | 25.3 ± 5.2 | N.R. | Unmedicated | MEGA-PRESS | 3 | water | Gannet 3.0 |
| ***Posterior MFC*** |  |  |  |  |  |  |  |  |  |  |
| Da Silva et al., 2019 (53) | 35 | 20.6 ± 1.6 | 18 | 21.3 ± 2.0 | 54.3 | Mixed | MEGA-PRESS | 3 | Water | Gannet |
| Modinos et al., 2018 (54) | 21 | 22.2 ± 3 | 20 | 23.7 ± 2.7 | 100.0 | Unmedicated | MEGA-PRESS | 3 | Cr | LCModel6.3-1L |
| Abbreviations: Cr, creatine; JRES, J-resolved; MEGA-PRESS, Mescher-Garwood point resolved spectroscopy; MFC, medial frontal cortex; SD, standard deviation; NR, not reported | | | | | | | | | | |

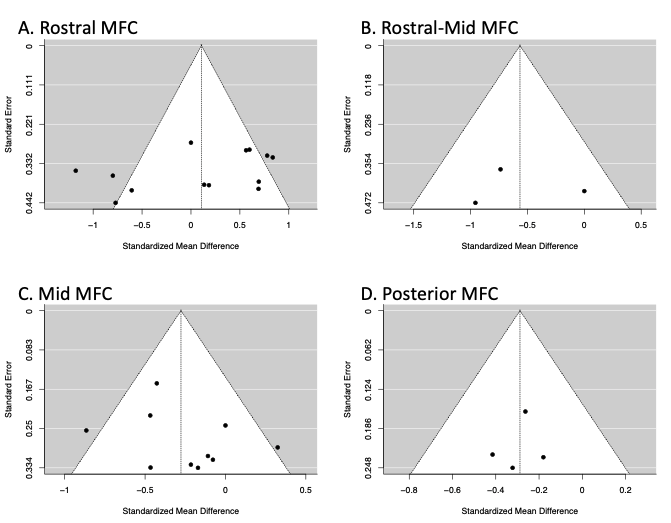

Supplementary Figure 1: Funnel plots for studies on Psychosis in the A) rostral MFC; B) rostral-mid MFC; C) mid MFC; and Posterior MFC showing the relationship between the standardized mean difference and standard error for each included study. Egger’s test is not significant for the rostral MFC (p = .14), rostral-mid MFC (p = .99), mid MFC (p = .31) or posterior MFC (p = .85).

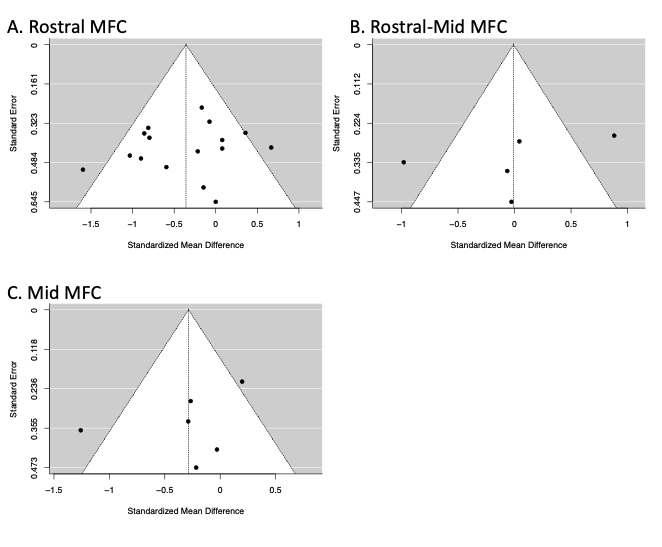

Supplementary Figure 2: Funnel plots for studies on depression in the A) rostral MFC; B) rostral-mid MFC; C) mid MFC, showing the relationship between the standardized mean difference and standard error for each included study. Egger’s test is not significant for the rostral MFC (p = .55), rostral-mid MFC (p = .44) or mid MFC (p = .50)

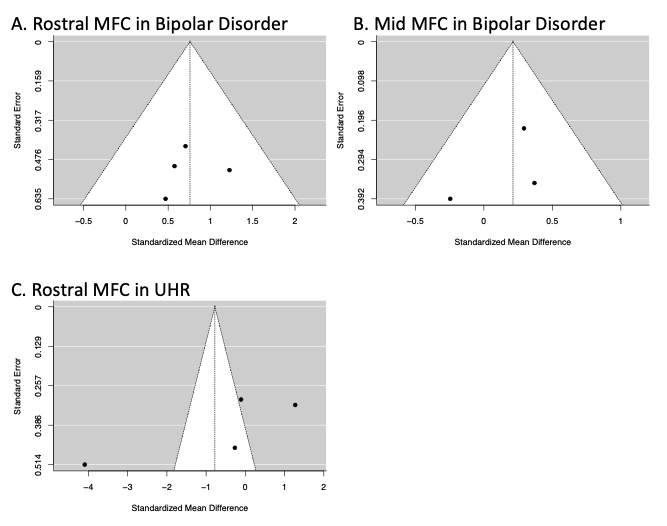

Supplementary Figure 3: Funnel plots for studies on bipolar disorder in the A) rostral MFC and B) mid MFC, and C, studies of individuals meeting ultra high-risk (UHR) criteria in the mid MFC, showing the relationship between the standardized mean difference and standard error for each included study. Egger’s test is not significant for the rostral MFC (p = .50) or mid MFC (p = .47) in bipolar disorder, or the rostral MFC in UHR (p = .06)

13. Bojesen KB, Broberg BV, Fagerlund B, Jessen K, Thomas MB, Sigvard A, *et al.* (2021): Associations Between Cognitive Function and Levels of Glutamatergic Metabolites and Gamma-Aminobutyric Acid in Antipsychotic-Naïve Patients With Schizophrenia or Psychosis. *Biol Psychiatry* 89: 278–287.

14. Brandt AS, Unschuld PG, Pradhan S, Lim IAL, Churchill G, Harris AD, *et al.* (2016): Age-related changes in anterior cingulate cortex glutamate in schizophrenia: A 1H MRS Study at 7 Tesla. *Schizophr Res* 172: 101–105.

15. Goto N, Yoshimura R, Moriya J, Kakeda S, Ueda N, Ikenouchi-Sugita A, *et al.* (2009): Reduction of brain γ-aminobutyric acid (GABA) concentrations in early-stage schizophrenia patients: 3T Proton MRS study. *Schizophrenia Research*, vol. 112. pp 192–193.

16. Öngür D, Prescot AP, McCarthy J, Cohen BM, Renshaw PF (2010): Elevated gamma-aminobutyric acid levels in chronic schizophrenia. *Biol Psychiatry* 68: 667–670.

17. Reid MA, Salibi N, White DM, Gawne TJ, Denney TS, Lahti AC (2019): 7T Proton Magnetic Resonance Spectroscopy of the Anterior Cingulate Cortex in First-Episode Schizophrenia. *Schizophr Bull* 45: 180–189.

18. Rowland LM, Krause BW, Wijtenburg SA, McMahon RP, Chiappelli J, Nugent KL, *et al.* (2016): Medial frontal GABA is lower in older schizophrenia: A MEGA-PRESS with macromolecule suppression study. *Mol Psychiatry* 21: 198–204.

19. Wang AM, Pradhan S, Coughlin JM, Trivedi A, Dubois SL, Crawford JL, *et al.* (2019): Assessing Brain Metabolism with 7-T Proton Magnetic Resonance Spectroscopy in Patients with First-Episode Psychosis. *JAMA Psychiatry* 76: 314–323.

20. Wijtenburg SA, Wang M, Korenic SA, Chen S, Barker PB, Rowland LM (2021): Metabolite Alterations in Adults With Schizophrenia, First Degree Relatives, and Healthy Controls: A Multi-Region 7T MRS Study. *Front Psychiatry* 12. https://doi.org/10.3389/fpsyt.2021.656459

21. Hjelmervik H, Craven AR, Sinceviciute I, Johnsen E, Kompus K, Bless JJ, *et al.* (2020): Intra-Regional Glu-GABA vs Inter-Regional Glu-Glu Imbalance: A 1H-MRS Study of the Neurochemistry of Auditory Verbal Hallucinations in Schizophrenia. *Schizophr Bull* 46: 633–642.

22. Marenco S, Meyer C, Kuo S, Van Der Veen JW, Shen J, DeJong K, *et al.* (2016): Prefrontal GABA levels measured with magnetic resonance spectroscopy in patients with psychosis and unaffected siblings. *American Journal of Psychiatry*, vol. 173 173: 527–534.

23. Tayoshi S, Nakataki M, Sumitani S, Taniguchi K, Shibuya-Tayoshi S, Numata S, *et al.* (2010): GABA concentration in schizophrenia patients and the effects of antipsychotic medication: A proton magnetic resonance spectroscopy study. *Schizophr Res* 117: 83–91.

24. Brennan BP, Admon R, Perriello C, LaFlamme EM, Athey AJ, Pizzagalli DA, *et al.* (2017): Acute change in anterior cingulate cortex GABA, but not glutamine/glutamate, mediates antidepressant response to citalopram. *Psychiatry Res - Neuroimaging* 269: 9–16.

25. Gabbay V, Mao X, Klein RG, Ely BA, Babb JS, Panzer AM, *et al.* (2012): Anterior cingulate cortex γ-aminobutyric acid in depressed adolescents: Relationship to anhedonia. *Arch Gen Psychiatry* 69: 139–149.

26. Gabbay V, Bradley KA, Mao X, Ostrover R, Kang G, Shungu DC (2017): Anterior cingulate cortex γ-aminobutyric acid deficits in youth with depression. *Transl Psychiatry* 7: e1216.

27. Hasler G, Neumeister A, Van Der Veen JW, Tumonis T, Bain EE, Shen J, *et al.* (2005): Normal prefrontal gamma-aminobutyric acid levels in remitted depressed subjects determined by proton magnetic resonance spectroscopy. *Biol Psychiatry* 58: 969–973.

28. Hasler G, Van Der Veen JW, Tumonis T, Meyers N, Shen J, Drevets WC (2007): Reduced prefrontal glutamate/glutamine and γ-aminobutyric acid levels in major depression determined using proton magnetic resonance spectroscopy. *Arch Gen Psychiatry* 64: 193–200.

29. Ironside M, Moser AD, Holsen LM, Zuo CS, Du F, Perlo S, *et al.* (2021): Reductions in rostral anterior cingulate GABA are associated with stress circuitry in females with major depression: a multimodal imaging investigation. *Neuropsychopharmacology* 46: 2188–2196.

30. Kantrowitz JT, Dong Z, Milak MS, Rashid R, Kegeles LS, Javitt DC, *et al.* (2021): Ventromedial prefrontal cortex/anterior cingulate cortex Glx, glutamate, and GABA levels in medication-free major depressive disorder. *Transl Psychiatry* 11. https://doi.org/10.1038/s41398-021-01541-1

31. Price RB, Shungu DC, Mao X, Nestadt P, Kelly C, Collins KA, *et al.* (2009): Amino Acid Neurotransmitters Assessed by Proton Magnetic Resonance Spectroscopy: Relationship to Treatment Resistance in Major Depressive Disorder. *Biol Psychiatry* 65: 792–800.

32. Walter M, Henning A, Grimm S, Schulte RF, Beck J, Dydak U, *et al.* (2009): The relationship between aberrant neuronal activation in the pregenual anterior cingulate, altered glutamatergic metabolism, and anhedonia in major depression. *Arch Gen Psychiatry* 66: 478–486.

33. Wang D, Wang X, Luo MT, Li YH, Wang H (2019): Gamma-aminobutyric acid levels in the anterior cingulate cortex of perimenopausal women with depression: A magnetic resonance spectroscopy study. *Front Neurosci* 13. https://doi.org/10.3389/fnins.2019.00785

34. Zhang X, Tang Y, Maletic-Savatic M, Sheng J, Zhang X, Zhu Y, *et al.* (2016): Altered neuronal spontaneous activity correlates with glutamate concentration in medial prefrontal cortex of major depressed females: An fMRI-MRS study. *J Affect Disord* 201: 153–161.

52. Wenneberg C, Nordentoft M, Rostrup E, Glenthøj LB, Bojesen KB, Fagerlund B, *et al.* (2020): Cerebral Glutamate and Gamma-Aminobutyric Acid Levels in Individuals at Ultra-high Risk for Psychosis and the Association With Clinical Symptoms and Cognition. *Biol Psychiatry Cogn Neurosci Neuroimaging* 5: 569–579.

53. Da Silva T, Hafizi S, Rusjan PM, Houle S, Wilson AA, Prce I, *et al.* (2019): GABA levels and TSPO expression in people at clinical high risk for psychosis and healthy volunteers: A PET-MRS study. *J Psychiatry Neurosci* 44: 111–119.

54. Modinos G, Şimşek F, Azis M, Bossong M, Bonoldi I, Samson C, *et al.* (2018): Prefrontal GABA levels, hippocampal resting perfusion and the risk of psychosis. *Neuropsychopharmacology* 43: 2652–2659.
